# Patient-reported adverse drug outcomes, from use of NSAIDs for chronic pain: a cross-sectional study in North India

**DOI:** 10.1101/2025.10.07.25337490

**Authors:** Kavita Bakshi, Ritu Tyagi, Garima Jaglan, Priyanka Rani, Acharya Balkrishna, Pratima Singh, Anurag Varshney

## Abstract

**Background:** Non-Steroidal Anti-inflammatory Drugs (NSAIDs) are commonly used to relieve pain and inflammation and are available over the counter in India. Despite their known adverse drug events (ADEs), formal reporting and patient awareness remain limited. This study aimed to evaluate the knowledge, attitude, and practices (KAP) of NSAIDs users, along with associated risks and self-reported ADEs, pivotal to developing strategies to mitigate the situation.

**Methods:** A cross-sectional study was conducted at a primary care facility in Haridwar, Uttarakhand, India. A structured questionnaire assessed KAP, risk factors, and ADE in 201 participants aged 18 to 75 years with continuous or intermittent NSAID use for ≥3 months for chronic pain.

**Results:** study outcomes revealed that 47% of subjects did not have proper knowledge of NSAIDS, 66% nursed a negative attitude, 55% were noncompliant to prescription, 63.20% were at risk for complications, and 66.66% developed ADEs. The data revealed that 68% of the study participants wanted to discontinue NSAIDs, whereas 58% expressed firm faith in the alternative and complementary systems for pain management.

**Conclusion and relevance:** The awareness about NSAID complications in the study participants was found to be limited in this specific geographical area. Most participants suffered from adverse drug events, were at risk, or required education. A record of participants showed dissatisfaction with NSAIDs, perhaps for higher cost, ineffectiveness, and concerns for safety. Maximum participants desired other treatment approaches with fewer or no side effects. Our study recommends a thorough review of existing guidelines on NSAID usage.

## 1. Introduction

Non-steroidal anti-inflammatory drugs (NSAIDs) are commonly used across the world for musculoskeletal pain relief and inflammation. World Health Organization (WHO) has cataloged NSAIDs on the list of essential medicines (Bindu et al., 2020). These are often used in relieving aches, inflammation, fever and, sometimes low-dose aspirin as an antiplatelet agent. NSAIDs are frequently prescribed in rural and urban areas of India as analgesics and antipyretics and are easily available over the counter without any prescription. This makes it difficult to conduct surveillance activities pertaining to NSAIDs. Health care facilities in India are rather inadequate; drugs are a bit expensive and not easily available to individuals living in remote areas and hilly regions. In order to save time and money, many participants tend to self-medicate regardless of NSAIDs causing adverse events and serious drug interactions at times (Moore et al., 2015).

Extensive pharmaco-epidemiological studies and meta-analyses of NSAIDs have described various health hazards especially gastrointestinal and cardiovascular complications arising from their prolonged usage. NSAIDs can affect different systems and damage vital organs (Bindu et al., 2020). The most common side effects are seen in the form of dyspepsia, diarrhea, constipation, peptic ulcer, acidity besides upper gastrointestinal bleeding. Dizziness, drowsiness, depression, headache, vertigo, and sleep disturbances due to reduced levels of melatonin and prostaglandins are believed to be linked to its side effects on central nervous system (CNS). NSAID-induced bronchospasm and asthmatic flares are well-known pulmonary side effects. In addition, an increase in blood pressure, precipitation of heart failure, anaphylactic shock and thrombosis are documented cardiovascular adverse effects of NSAIDs (Schjerning et al., 2020). In addition, NSAID can elevate aminotransferases levels and cause fulminant hepatic failure. NSAID-induced simple skin irritation to Steven Johnson syndrome is well-known adverse event of the integumentary system (Ye et al., 2019).

NSAIDs can increase complications in participants with hypertension, diabetes mellitus, coronary artery disease, bronchial asthma, stroke, liver disease, and chronic kidney diseases. It can interact with selective serotonin reuptake inhibitors, corticosteroids, antiplatelet and anti-thrombotic medications. NSAIDs in the geriatric population enhance the risk of side effects and drug interactions owing to the versatility of polypharmacy (Chi et al., 2018)(Gnjidic et al., 2014). Considering these facts, increased mortality and morbidity due to NSAIDs has been implicated in the elderly (Ho et al., 2020)(Oliva et al., 2022), however, no study has so far described the attitude, risks and side effects of NSAIDs usage in the Indian population.

More specifically, previous studies have focused on NSAID-induced adverse drug events (ADE) in orthopedic patients (Gor & Saksena, 2011). In another study carried out by Chatterjee, the prevalence of NSAID-induced gastrointestinal, renal, and cardiovascular complications was also documented (Chatterjee et al., 2015). However, none of the above-mentioned studies focused on patient Knowledge, Attitude, and Practices (KAP) related to the consumption of NSAIDs. Patient KAP is an important aspect that leads to the identification of drug use patterns, perceptions and shortcomings that require correction through further research and policies. Therefore, we planned to identify the specific groups of patient population requiring more education, and care with NSAIDs use. The current study was therefore designed to explore KAP, risk factors and ADE among NSAID users with chronic pain conditions. The results of this study can impact current policies and guidelines for the use of NSAID therapeutics in India. Furthermore, the study findings also highlight the need for advancement of research related to NSAIDs.

## 2. Materials and Methods

### 2.1 Sample and settings

This was a cross-sectional observational study on individuals visiting Patanjali Ayurveda Hospital, Haridwar, Uttarakhand, India between December 2022 to February 2023. The ethical approval for the study was obtained from the Institutional Ethics Committee-Patanjali Bhartiya Ayurvigyan Evam Anusandhan Sansthan, Haridwar, (vide approval number-PAC/IEC/2022/36). Adult individuals, aged 18-75 years, and on regular or intermittent use of NSAIDs for ≥ 3 months, for pain related complaints, were enrolled in the study after taking informed consent from each participant. The STROBE statement was followed for the entire study. A structured case record form was used for data collection. Skilled interviewers collected data after providing comprehensive details to the participants regarding the study, its purpose, and the voluntary nature of participation. Furthermore, the participants were assured that all information would be kept confidential and anonymous, along with their right to discontinue participation at any time. Prior to the data collection, written informed consent was obtained from each participant.

#### Questionnaire and its administration

A team of physicians, pharmacists, and scientists reviewed the questionnaires to ensure accuracy. Unrelated questions were eliminated or replaced, to align with study objectives. Subsequently, 20 individuals were invited to verify the understanding, readability, and compatibility of the questionnaire with the study objectives. A case record form was used to collect data from each participant. The first section of the case record form contained information on the demographic characteristics, medical history and social history of the individual. The second section consisted of questions probing knowledge, attitudes, and practices. There were five questions on knowledge, 8 questions of attitude and six on practice. Each question had three possible answers. “No,” “do not know,” and “yes” were the possible responses to the knowledge questions, while “no,” “not sure,” and “yes” were the responses to the attitude questions. Knowledge-related questions focused on the awareness of risk, interaction, disease precipitation, dependence, and the presence of healing properties. The first four attitude-related questions covered pain alleviation, cost-effectiveness, satisfaction, and the desire to continue therapy. The final four questions inquired about receptivity to different treatments, whether allopathic or alternative, and general beliefs in alternative medicine. There were various options for each practice-related question, which were then further divided into categories of good, bad, and not so good practices. The first question asked how long the drug had been used, with options of 3 months to 2 years, 2-5 years, and > 5 years. Other options included 1-3 times per week, 4-7 times per week, and > 7 times per week to score the frequency of current drug use. Prescription modalities were also recorded with options including medical doctors, pharmacists, and self-prescriptions. Dosage was assessed based on whether it was in accordance with the prescription, exceeded the prescription, or whether the patient was uninformed about the dosage. The use of NSAIDs and antacid consumption was also probed, with the response options being yes, no, or uninformed. The last question pertained to the use of other drugs before, after, or at any point. For practice-related questions, each of the aforementioned possibilities received ratings ranging from good, bad, and severe respectively. The third section of the questionnaire discussed the adverse effects of the medication on various organ systems, based on the symptoms described by individuals. According to the NICE guidelines, associated risk factors such as anticoagulants, corticosteroids, aspirin, SSRI, comorbidities, and conditions that may enhance NSAID-induced concerns have been listed (*NICE,* 2018). The data collection form was developed and approved by specialists.

### 2.2 Statistical Analysis

The analysis was performed using Microsoft Excel and IBM SPSS software (version 26.0). Categorical data were explained in terms of frequencies and percentages. Continuous data was expressed as mean and standard deviation (SD). Pearsons’s correlation coefficient was utilized to determine correlations among variables. The minimum required sample size was determined to be 186 using Raosoft (http://www.raosoft.com), with a margin of error of 5%, confidence level of 95%, population size of 3600, and a response distribution of 85%. This computation was based on a cross-sectional study on the prevalence of NSAID-induced complications in India (Chatterjee et al., 2015).

The questionnaire has a Cronbach’s α value of 0.85, indicating good reliability. All the answers were marked from 1 to 3, from more desirable to less desirable. Each answer of the section was added to provide a total. The good and bad categories of KAP score were determined based on the median value of the total score. Ethical principles listed in the Declaration of Helsinki and all applicable laws and regulations, including data privacy laws, were strictly followed.

## 3. Results

A total of 273 individuals were screened for chronic pain at a hospital in Haridwar, India. These included individuals who had already received pharmacological treatments. Of these, 218 individuals, aged between 18 and 75 years, fulfilled the inclusion criteria for the use of NSAIDs for chronic pain. Later, 17 individuals were excluded from the study due to missing information in ‘case record form’ and the data of 201 individuals was analyzed.

### 3.1 Demography

The average age of the participants was found to be 50.08± 13.79 (***Table 1***) of which 79.10% of females outnumbered 20.90% of male participants. The study consisted of 89.10% of participants that were married and 10.90% who were unmarried. Most of the study participants (54.73%) had attended school and 45.27% had attended college. In this study, consumption of alcohol and tobacco was found to be as low as 13.90%, and 12.90% respectively. Most participants (77.10%) followed a vegetarian diet. Socio-economic categories were derived from the Kuppuswamy scale (Sood & Bindra, 2022). This revealed that 10.00% of participants belonged to the ‘upper class,’ 50.20% to the ‘upper middle class,’ 32.30% to the lower middle class, 7% to the ‘upper lower’ and 0.5% belonged to the ‘lower’ socio-economic background. In addition to chronic pain, study participants reported several other co-morbid conditions. Among these, hypertension (25.87%) was the leading cause followed by hypothyroidism (17.41%) and diabetes mellitus (16.90%); there were 1.49% individuals nursing a chronic kidney disease, liver disease, or varicose veins. In this study, 2.98% of participants with bronchial asthma and 01.99% suffering from coronary artery disease and 0.50% having anemia were also documented.

**Table 1.** Demographic details of the participants along with comorbidities.

| Characteristics of the population |  | Total N (%) |
| --- | --- | --- |
| Age (in years) |  | 50.08 ± 13.79* |
| Gender | Male | 42 (20.90) |
|  | Female | 159 (79.10) |
| Marital status | Married | 179 (89.10) |
|  | Unmarried | 22 (10.90) |
| Education | College | 91 (45.27) |
|  | School | 110 (54.73) |
| Alcohol | Yes | 28 (13.90) |
|  | No | 173 (86.10) |
| Tobacco | Yes | 26 (12.90) |
|  | No | 175 (87.10) |
| Diet | Vegetarian | 155 (77.10) |
|  | Nonvegetarian | 46 (22.90) |
| Monthly Family income (INR) | >185,895 | 39 (19.40) |
|  | 92951-185894 | 38 (18.90) |
|  | 69535-92950 | 20 (10.00) |
|  | 46475-69534 | 56 (27.90) |
|  | 27883-46474 | 17 (08.50) |
|  | 9308-27882 | 14 (07.00) |
|  | <9307 | 17 (08.50) |
| Socio economic category | Upper class | 20 (10.00) |
|  | Upper middle class | 101 (50.20) |
|  | Lower middle class | 65 (32.30) |
|  | Upper Lower | 14 (07.00) |
|  | Lower | 01 (00.50) |
| Comorbidities | Hypertension | 52 (25.87) |
|  | Diabetes Mellitus | 34 (16.90) |
|  | Hypothyroidism | 35 (17.41) |
|  | Coronary Artery Disease | 04 (01.99) |
|  | Asthma | 06 (02.98) |
|  | Chronic Kidney Disease | 03 (01.49) |
|  | Liver Disease | 03 (01.49) |
|  | Anemia | 01 (00.50) |
|  | Varicose Vein | 03 (01.49) |

\*Standard Deviation, total individuals participated =201

### 3.2 Pain conditions and KAP

The participants using NSAIDs are shown in **Figure 1 A**. The reasons for using this drug include knee osteoarthritis, headache, rheumatoid arthritis, and other painful musculoskeletal conditions. The study revealed that very few people indicated toothaches as a reason for using medicine. **Figure 1 B** shows that the knowledge of the respondents about NSAIDS was inadequate. There were five questions to estimate the knowledge of the individual. As per the study, 26% perceived no risks associated with use of NSAIDs, 16% were unsure, and 58% individuals were aware of the risks associated with NSAIDs consumption. About 49% were aware of chances of potential drug interactions, 22% were unsure, and 29% assumed there were no interactions. Of all 59% respondents were aware that NSAIDs could precipitate diseases, 22% were uncertain, and 19% held an erroneous belief that it could not. When inquired about dependence on NSAIDs, 43% opined that one could become dependent, while 9% were not sure and 48% knew it was not habit forming. While we inquired about healing properties of NSAIDS, 35% had a incorrect notion that NSAIDs could heal, 15% were uncertain and 50% agreed it did not have healing properties. Briefly, 47% lacked proper knowledge of NSAIDs and required education.

**Figure 1.**
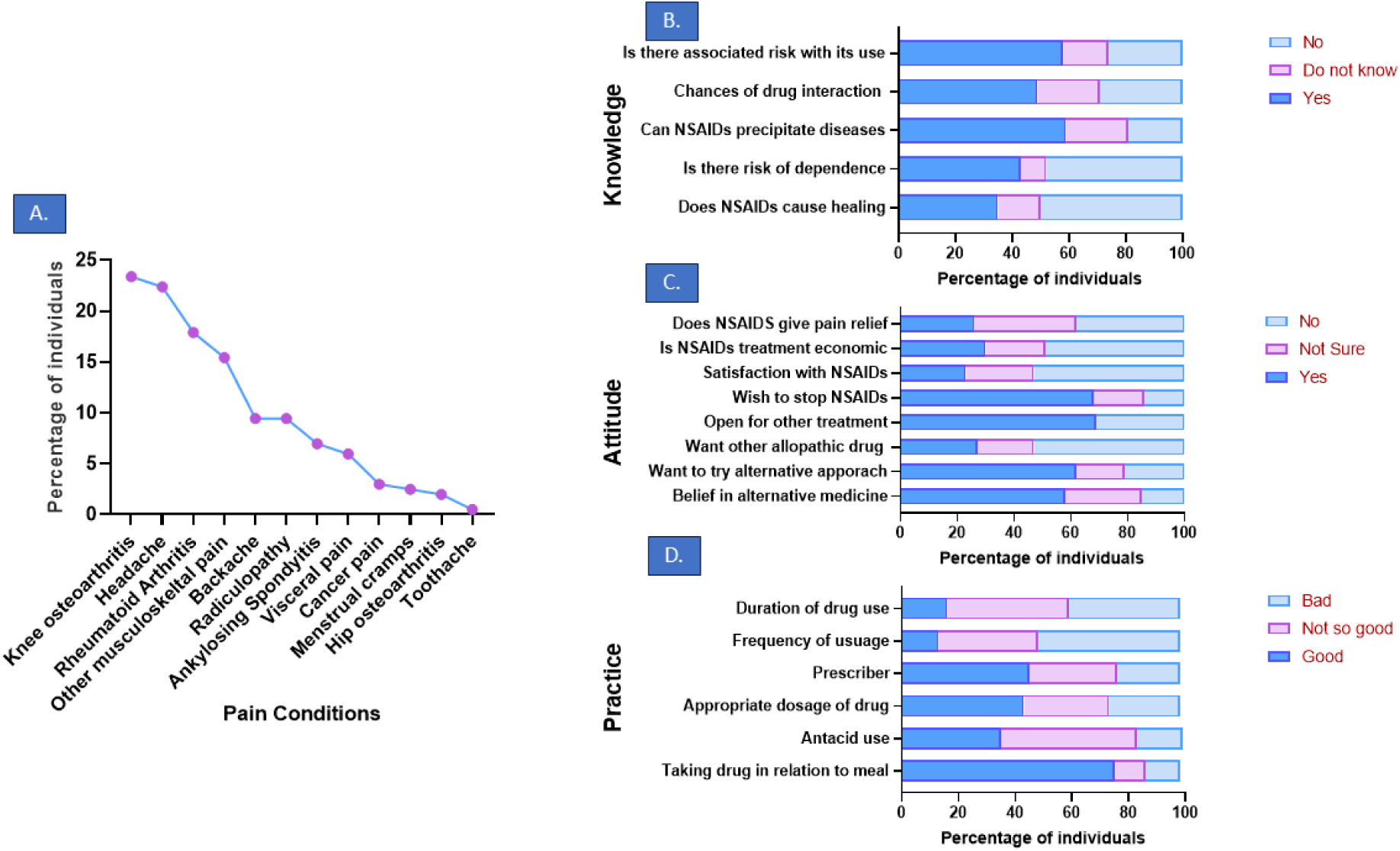
A. Pain conditions; B. Current knowledge; C. Attitude; D. Practices followed amongst patients taking NSAIDs for pain relief.

There were eight questions related to attitude; the first four asked about their perception towards NSAIDs and the last four asked if the person wanted to switch to another treatment (**Figure 1 C)**. There were 26% individuals who agreed NSAIDs gave them relief from pain, 38% refused, and 36% provided a mixed response. Thirty percent of respondents considered NSAIDs to be economical, 21% were uncertain, and 49% considered it expensive. Satisfaction with NSAIDs was low as 53% showed clear dissatisfaction with NSAIDs while 23% were satisfied, and 24% were unsure about it. When asked if they wanted to stop current treatment, 68% affirmed yes, 18% were not sure and 14% wanted to continue current treatment. Survey responses showed that 69% of participants wanted to try another treatment while only 31% said they were not open to exploring another treatment regimen. It is interesting to note that 53% refused to try another drug for pain relief and only 27% were ready for it, while 20% were uncertain of it. However, when asked if they were ready for an alternative and complimentary medicine, 62% agreed, 21% disagreed, and 17% were uncertain. Majority of the respondents (58%) showed firm belief in alternative and complementary medicine, while 15% disagreed and 27% were unsure of its benefits. Briefly, three quarters of population (66%) held a negative attitude towards NSAIDs and only 34% showed positive attitude towards NSAIDs.

Figure 1 **D** shows the responses to practice-related questions on NSAIDs use. There were 16 % respondents who were taking NSAIDs for less than two years, 44% from two to ten years, and 40% for more than ten years. A pattern of frequent NSAID use was also observed. The frequency of taking the drug in last month of up to <4 times a month accounted for 14%, 2-4 times a week for 36% and > 4 times a week for 50% of the participants. Prescribing trends accounted for 46% by doctors, 32% by pharmacists and 22% of self-referrals. As many as 44% agreed that they were taking appropriate dose, 31% were taking more than prescribed and 25% responded that they were uncertain, implicating a divergence from good practices. In this study, 35% of individuals strictly adhered to prescribed antacids or proton pump inhibitors along with NSAIDs, 48% were noncompliant., and 16% were ignorant. When asked about the intake of NSAIDs related to meal a total of 75% confirmed they took it post prandial, 11% took before meal and 12% took anytime during the day. Briefly, 55% of respondents had adopted bad practices, while 45% were following good practices regarding the use of NSAIDs.

### 3.3 Adverse Drug Events

NSAIDs used for long periods of time have been associated with adverse effects. In the present study, the gastric system was impacted the most, with 30.85% individuals complaining of it, followed by an impact on nervous and integumentary systems with 8.96% and 7.46% respectively (Figure 2 **A**). Percentages of individuals with complaints related to kidneys, respiratory system and cardiac system were 4.98%, 3.48% and 3.48% respectively. When the adverse effects associated with drug use were evaluated, we found that paracetamol, diclofenac and ibuprofen, showed the maximum adverse events (Figure 2 **B**). Celecoxib and piroxicam showed minimal impact, whereas doloxicam and ketorolac showed no adverse impact among the participants. A total of 43.78% of individuals were taking a combination of paracetamol and other NSAID, which has shown maximum number of Adverse Drug Events (ADE).

**Figure 2.**
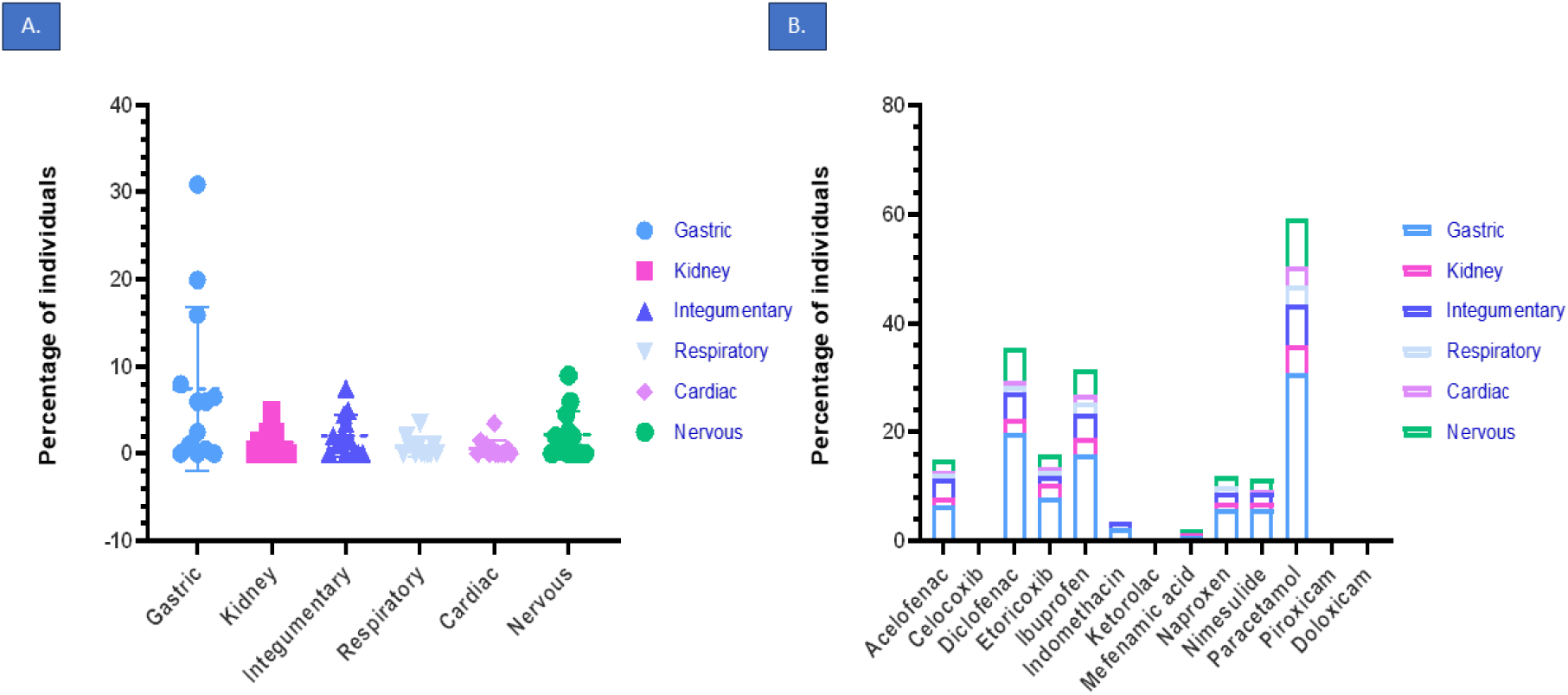
A. Adverse drug events (ADE) reported by individuals impacting various systems, B. Drugs taken for pain relief along with ADE

NSAID-related adverse events were noted in a total of 66.66% individuals. Among them, 97.03% of participants cumulatively experienced gastrointestinal side effects, such as dyspepsia, upper abdominal pain, and acidity. CNS-related side effects like dizziness, headache, sleep disturbance and reduced memory were found to be affected in 28.37% participants. Integumentary system-related adverse events like skin rashes, hair loss and pigmentation were noted in 26.87% participants. The lesser noted NSAID-related side effects were recorded in the renal system with reduced urination, urgency, dribbling, burning sensation while urinating, pain in flank, and or ankle edema in 17.41%; the respiratory system was found affected concomitant with breathing issues, cough, nasal congestion among the 9.45 % individuals; the cardiovascular system related problems like altered blood pressure, temple pain, nose bleeding, and chest pain were observed in 7.47% participants respectively. In addition, there were 46 (22.38%) participants having adverse drug effects involving more than one organ system.

### 3.4 Risk Factors

According to “NICE’s Practice Advisory on Appropriate Use of NSAIDs in Primary Care’’, stated in Clinical Knowledge Summaries, 63.20% individuals were at risk of NSAID use (***Table 2***). The risks included the presence of certain comorbidities, such as old age, lifestyle, and other interactive drugs. Approximately 25.90% of participants were reported to be hypertensive, indicating the highest number among all other comorbidities followed by 17.40% with hypothyroidism and 16.90% with diabetes mellitus. A few participants (10.94%) were reportedly taking corticosteroids and antiplatelet drugs (5.5%), along with NSAIDs, which are known to predispose one to vascular events like gastrointestinal bleeding (Ho et al., 2020). There were 14.93% individuals above age 65 consuming the drug, which increases their risks for peptic ulcers, cardiovascular events, and renal events (Marcum & Hanlon, 2010). A total of 127 individuals were at risk of NSAIDs use, along with 64 individuals with multiple risk factors.

**Table 2.** Risk factor* analysis in NSAIDs users with chronic pain.

| **Risk Factors** | **Individual (%)** |
| --- | --- |
| On anti-coagulants/blood thinners like heparin | 02 (00.99) |
| On antiplatelet drugs like aspirin | 11 (05.50) |
| On corticosteroids | 22 (10.94) |
| On selective serotonin reuptake inhibitors | 01 (00.49) |
| History of hypertension | 52 (25.90) |
| History of coronary artery disease/myocardial Infarctions | 04 (02.00) |
| History of diabetes mellitus | 34 (16.90) |
| Age above 65 years | 30 (14.93) |
| History of asthma | 06 (03.00) |
| History of chronic kidney disease | 03 (01.49) |
| History of liver disease | 03 (01.49) |
| History of hypothyroidism | 35 (17.40) |
| Anemia | 21 (10.40) |
| Total Individuals with risk | 127 (63.20) |

\*NICE Clinical Knowledge Summaries: NSAIDs - prescribing issues revised August 2018 https://cks.nice.org.uk/nsaids-prescribing-issues

### 3.5 Correlation between variables

The correlation among knowledge, attitude, practices, risk factors, ADEs and various demographic factors are tabulated (***Table 3*)**. Interestingly, the risk factors showed a positive correlation with age and a negative correlation with income. Individuals with prolonged pain conditions showed a positive improvement in KAP. Education and higher income showed a positive relationship with good knowledge, and lower socioeconomic status was correlated with improper practices.

**Table 3.**
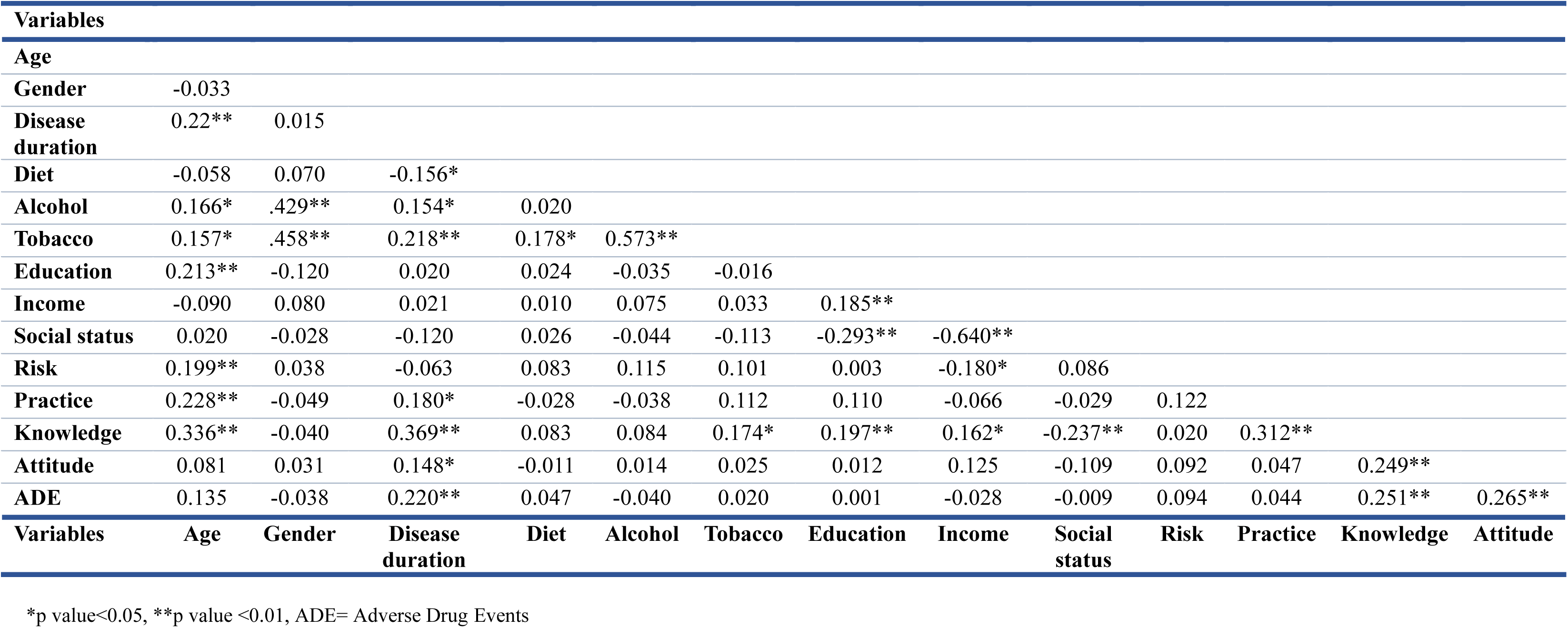
Correlation matrix for variables.

## 4. Discussion

This is the first study from India to indicate a growing demand for alternative and complementary systems for the management of chronic pain in addition to providing NSAID-induced adverse events along with risk and KAP. By providing a unique opportunity to analyze the effects of NSAIDs on the population lacking adequate knowledge and compliance towards NSAIDs usage highlighting a need to improve the prescription patterns including self-medication in India.

The results unambiguously revealed that half of the population lacked adequate knowledge and was noncompliant. Almost two-thirds of the population wanted to discontinue NSAIDs. In order to maintain safe and effective use of NSAIDs, it is imperative to take into account an individual’s knowledge, attitudes, and habits (Sinuraya et al., 2023). Lack of knowledge, time restrictions, unpredictability, and insufficient training in Adverse Drug Reaction (ADR) diagnosis remain major barriers to the inadequate reporting of NSAID-related adverse events (ADE) in India (Dash et al., 2023). ADE reporting is a complex and time-consuming process but becomes easier, faster, and more effective when patients report themselves (Pelletier et al., 2016). Current study highlights the incidence of patient-reported NSAID-induced adverse drug events seen in 66.66% of the study population, with 63.20% having risk for complications. The increased dissatisfaction expressed in the responses indicates an interest in alternative and complementary medicine.

### 4.1 Knowledge Attitude and Practices (KAP)

Our findings reveal notable gaps in knowledge, with over half of participants unaware of potential drug-drug interactions despite 59% acknowledging the risks of NSAIDs use. Similar reports from other populations highlight that inadequate awareness contributes to inappropriate use and risk of adverse outcomes, underscoring the need for targeted patient education (Arain et al., 2019). Attitudes towards NSAIDs were ambivalent, with 53% of participants dissatisfied due to adverse effects and higher cost. A significant proportion (62%) expressed openness to alternative or complementary therapies, and 58% reported strong confidence in their efficacy. This aligns with global trends where patients increasingly seek integrative approaches, such as yoga and traditional medicine, to manage chronic pain while minimizing adverse drug events (Basedow et al., 2014)(Bakshi et al., 2025). Such preferences are consistent with international clinical guidelines that recommend non-pharmacological and mind-body practices as adjuncts in knee osteoarthritis management (Brosseau et al., 2017). About practices, one-third of participants reported consuming higher-than-recommended NSAIDs dosages, and 11% used them on an empty stomach. Only 35% were taking prescribed concomitant gastroprotective agents, leaving a large proportion at risk of gastrointestinal complications. These findings are consistent with international reports of suboptimal NSAID practices and reflect the challenges of self-medication, particularly in low- and middle-income settings (Sulaiman et al., 2012). Collectively, our findings highlight the need to enhance patient awareness, promote rational prescribing, and integrate safe complementary approaches to optimize pain management, with particular attention to individuals from lower socioeconomic groups to improve their knowledge, attitudes, and practices.

### 4.2 Adverse Drug Events (ADEs)

Although NSAID use in our study was predominantly intermittent, the incidence of ADEs remained high, with 66.6% of participants reporting at least one event. Gastrointestinal complications were most frequent (30.8%), presenting as acidity, dyspepsia, abdominal pain, and constipation. This aligns with previous reports identifying gastrointestinal problems as the predominant adverse effect of NSAIDs (Jarernsiripornkul et al., 2009). The underlying mechanism involves cyclooxygenase (COX) inhibition, which reduces prostaglandin synthesis essential for mucosal protection, thereby predisposing patients to gastrointestinal bleeding and ulceration (McEvoy et al., 2021)

Other organ systems were also affected, albeit at lower frequencies. Central nervous system complaints (8.9%) included dizziness and headaches, while dermatological reactions (7.5%) involved hair loss, pigmentation, and rashes, consistent with prior reports (Lopes et al., 2018). Cardiovascular complications (3.5%) such as elevated blood pressure, palpitations, and angina were less common but clinically significant, given that diclofenac and ibuprofen, the most widely used NSAIDs are known to elevate cardiovascular risk (Ruschitzka et al., 2017). Paracetamol, often perceived as safer, was also implicated in gastrointestinal and cardiovascular events, particularly when combined with NSAIDs, which doubles the risk of gastric bleeding (Richard et al., 2023).

Taken together, our findings highlight the multifaceted ADE profile of NSAIDs across gastrointestinal, cardiovascular, renal, and neurological systems. Growing evidence suggests that high-dose diclofenac carries risks comparable to rofecoxib, which has been withdrawn from the market, raising concerns about its continued inclusion in essential drug lists (McGettigan & Henry, 2013). Safer prescribing practices, dose minimization, and the use of topical formulations where possible are therefore critical to reducing the burden of ADEs in populations requiring long-term pain management.

### 4.3 Risk Factor

Risk factors significantly influence the likelihood of NSAID-related complications. In our study, 31.8% of participants were identified as being at risk, particularly those of advanced age. Current clinical guidance emphasizes that NSAID therapy should be individualized, with risk-benefit assessments conducted prior to initiation and patients closely monitored throughout treatment (Ho et al., 2020). An interesting study showed a similar finding, wherein 51% of patient population was prescribed NSAIDs even when at risk or when not recommended thereby compounding the potential for harm (Lanas et al., 2011). NSAID use has been associated with gastrointestinal, renal, and cardiovascular complications, and chronic paracetamol consumption may further aggravate blood pressure control (Abbasi & Teakell, 2023). Long-term and frequent analgesic use has also been linked to functional decline and increased morbidity in older adults (Andersen et al., 2023). These findings highlight the importance of considering comorbidities, age, polypharmacy, and concomitant risk factors when prescribing analgesics. Overall, our results reinforce the need for judicious prescribing and careful monitoring to minimize preventable adverse outcomes. Strengthening deprescribing strategies, particularly in high-risk and geriatric populations, remains an important public health priority.

### 4.4 Limitations of the study

The present study had a predominance of female participants using NSAIDs under medical supervision. Given the widespread over the counter and unsupervised use of NSAIDs, these findings may not be fully generalizable to the broader population. As a cross-sectional design was employed, causal relationships cannot be established and recall bias may have contributed to missing or under-reported information.

### 4.5 Recommendations

A study with a larger sample size should be conducted to improve the power of statistical analysis. Multicentric cohort studies may help reduce bias and improve generalizability. Patient education programs should be encouraged to enhance knowledge about the use of their drugs with patient information leaflets, regular community-based education programs to reduce possible complications, and proper usage of NSAIDs. The study findings will be of immense use to Central Drugs Standard Control Organization (CDSCO) to contribute to the guidelines and recommendation of NSAIDs use in India.

## 5. Conclusion

In this study, most participants were on long-term use of NSAIDs and many of them may have nursed negative attitudes towards the drugs, probably owing to dissatisfaction with their efficacy, higher costs and ADE. About one-third were at risk, and more than half had already experienced ADE. NSAID-induced systemic side effects seem to further deteriorate general health evidence from the responses obtained. Some participants held incorrect beliefs and practices related to NSAIDs and required education. The participants showed openness to newer and better treatment modalities, which can facilitate integrating complementary and alternative health practices that are believed to be safer and acceptable. We propose establishing and implementing guidelines to deprescribe NSAIDs to high-risk individuals to control ADE and reduce the burden on the healthcare system.

## Glossary

NSAIDs: Non-Steroidal Anti-Inflammatory Drugs
WHO: World Health Organization
CNS: Central Nervous System
ADE: Adverse Drug Events
KAP: Knowledge Attitude and Practices
STROBE: Strengthening The Reporting of Observational Studies in Epidemiology
ADR: Adverse Drug Reaction
COX: Cyclooxygenase
CDSCO: Central Drugs Standard Control Organization

## Human Ethics and Consent to Participate declarations

The ethical approval for the study was obtained from the Institutional Ethics Committee of Patanjali Bhartiya Ayurvigyan Evam Anusandhan Sansthan, Haridwar, Uttarakhand, India (vide approval number-PAC/IEC/2022/36). All participants provided written informed consent before enrolling into the study.

## Data Availability

All data produced in the present study are available upon reasonable request to the authors

## Acknowledgment

We sincerely thank Patanjali Ayurveda Hospital for their support in enrolling the participants at the study site. We thank Dr. Robindranath Das and Dr. Himanshu Sharma for reviewing the manuscript. We express our gratitude to the participants for taking time and energy to contribute to this research.

## Author Contributions

Kavita Bakshi, Pratima Singh, and Anurag Varshney were accountable for the conceptualization and design of the study. Kavita Bakshi, Ritu Tyagi, Garima Jaglan, and Priyanka Rani collected data. Kavita Bakshi analyzed and visualized the data under the supervision of Anurag Varshney and Acharya Balkrishna. Kavita Bakshi wrote the first draft of the manuscript and incorporated changes after a review by Ritu Tyagi, Garima, and Priyanka Rani. Anurag Varshney and Acharya Balkrishna held responsibilities for the project study administration. All the authors have read the manuscript and approve of it.

